# Strong Public Disapproval of Proposed Reductions in Psychologist Competency Standards in Ontario: Findings from 1,141 General Community Adults in an Observational Cohort Study

**DOI:** 10.1101/2025.11.21.25340610

**Authors:** James MacKillop, Emily MacKillop, Kyla Belisario, Ilvy Goossens, Sabrina K. Syan, Brian M. Bird, Danielle Rice, Taylor Hatchard, Emily E. Levitt

## Abstract

The regulatory body for psychologists in Ontario, the College of Psychologists and Behavior Analysts of Ontario, has proposed sweeping reductions in competency requirements. These include elimination of the doctoral degree requirement, dramatic reductions in clinical training, and elimination of specializations and would make Ontario the largest jurisdiction with the lowest competency standards in North America. There has been considerable public opposition from active practitioners, but public attitudes toward these changes are unknown. The current study assessed attitudes in 1,141 general community adults in Ontario (mean age = 42.7, 62.1% female) in an ongoing longitudinal observational cohort study. The large majority disapproved of the proposed changes (71%), whereas 20% reported no opinion and 9% approved of the changes. The modal response was strong disapproval (44%), which was more than half of those who disapproved, whereas among those who approved, only a small proportion expressed strong approval (2%). In order of frequency, reasons for disapproval were reduced quality of healthcare (94%), more healthcare providers with a lower skillset and competencies (85%), higher probability of misdiagnosis (78%), and increased risk in high-stakes contexts (69%), increased likelihood of conflicting medical opinions (55%), and adverse impacts on the current workforce and trainees (both 54%). When asked about the single most important concern, 62% identified reduced quality of healthcare followed by an increased number of healthcare providers with lower professional competency (22%). Responses to open text options were highly consistent with the quantitative results (e.g., “*We don’t need fewer professional standards--we need highly skilled professionals to deal with an increasingly complex world.”*). Collectively, the results suggest strong disapproval for the proposed reductions in psychologist competency standards among general community adults in Ontario.

**VISUAL ABSTRACT:** Simplified Distribution of Public Attitudes toward Psychologist Competency Reductions (n=1,141)

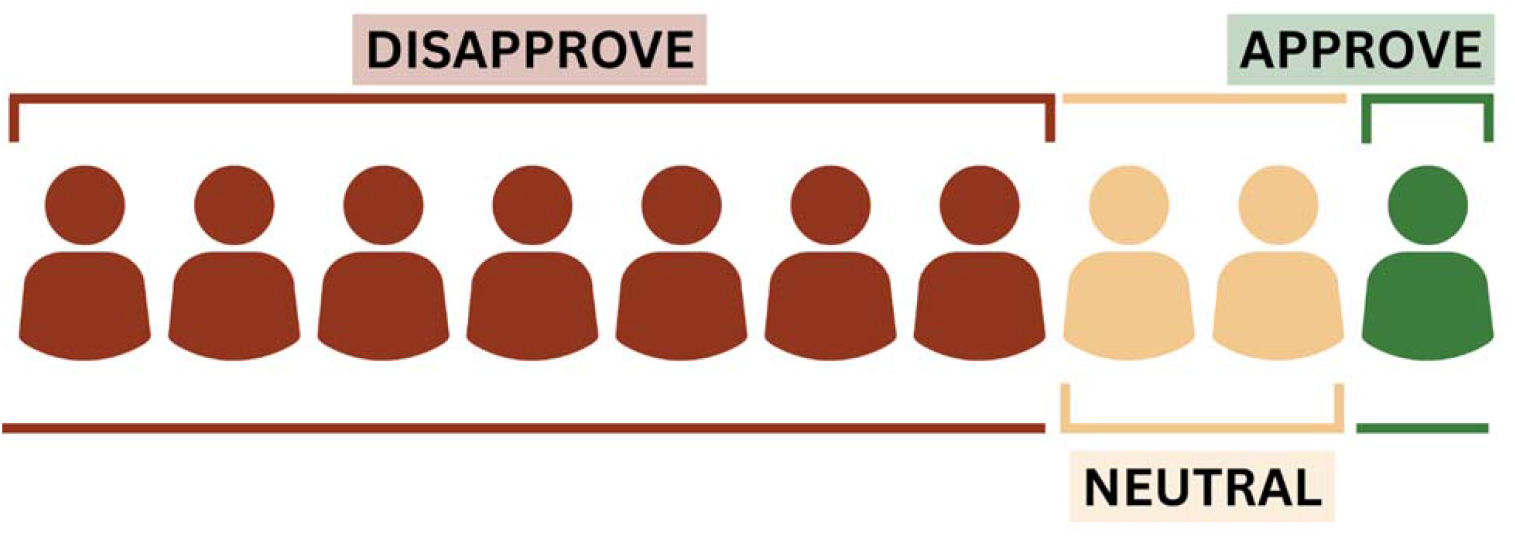

## INTRODUCTION

Psychologists are specialized mental health providers who conduct complex assessments, provide evidence-based behavioural interventions, and consult extensively with other healthcare workers across sectors. Psychologists are extensively trained for these roles, and their clinical decisions are determinative for quality healthcare, access to services, and public safety. Insurers, health authorities, and physicians often recognize psychologists as professionals who are uniquely qualified to conduct standardized assessments, interpret complex data, and integrate findings across cognitive, emotional, and behavioural domains.

Given the consequential roles in complex and high-stakes healthcare contexts, the training to become a psychologist is rigorous. The field standard in North America includes completion of an accredited doctoral degree (PhD, PsyD, EdD), with substantial hands-on clinical training (including both practicum exposures and a one-year full-time predoctoral residency); supervised postdoctoral practice; and completion of three examinations (i.e., the Examination for Professional Practice in Psychology [EPPP], a standardized exam on knowledge of the field; a jurisdiction-specific jurisprudence examination; and an oral examination by existing practitioners) (1–3). These requirements aim to ensure that clinical psychologists receive the training required to be appropriately skilled in their scope of practice. Extensive as it is, however, it reflects a baseline but not the endpoint of training. Because of the clinical complexity psychologists often face, many seek additional advanced training to achieve board certification in specialty areas, such as neuropsychology, forensic psychology, and school psychology (4).

These are the predominant current standards in Ontario, Canada. To further protect the public, eight specializations exist in Ontario (5), intended to ensure depth of expertise and quality of care across diverse mental health needs. Psychologists are registered to practice specifically within one or more of these specializations to ensure they practice competently. Further, identified scopes of practice are intended to signal to the public that individuals are proficient in providing specialized care in specific domains. In other words, not every psychologist is anticipated to be fully competent for every clinical population or every context, similar to subspecialties in medicine.

On September 26, 2025, council members of the regulatory body for psychologists in Ontario, the College of Psychologists and Behavioral Analysts of Ontario (CPBAO), passed a motion to substantially reduce the psychologist competency requirements in Ontario (6,7). Broadly speaking, six major changes were proposed. The first was removing the requirement for a doctoral degree (PhD, PsyD, EdD), reducing the formal coursework and training in research and evidence appraisal, and eliminating the one year 1600-2000 hour predoctoral clinical residency year. Similarly, the second was reducing the minimum number of practicum placements prior to the residency, from 3 or more (∼2000 hrs) to a minimum of 1 practicum (∼450-850 hours). The third was permitting unlimited attempts of the EPPP examination, the aforementioned exam of a candidate’s foundational knowledge in psychology and judgment in matters of general jurisprudence and ethics required for registration as a psychologist in the United States and Canada. The fourth was replacing the Ontario-specific Jurisprudence and Ethics Examination (JEE) with an online “low-stakes” ethics webinar with a “no-fail” examination (8). The fifth was removing the requirement for an oral examination. The sixth was removing the regulation of patient age groups (e.g., children, adults, seniors) and scopes of practice (e.g., clinical; clinical neuropsychology; forensic/correctional; health; counselling, industrial/organizational; school; rehabilitation), collapsing the eight practice areas into two broad categories: health service and industrial/organizational. Additional nuances included allowing accrediting bodies outside Canada and the United States and a consolidation of titles.

In the broader context of healthcare challenges in Ontario, the proposed reductions in competency standards are putatively to increase the number of psychologists in the province, thereby improving access and healthcare; to respond to new the legislation requiring timely credentials review; and to address potential concerns from the Office of Fairness Commissioner. A close analysis of these pretexts is beyond the current scope, but, given the dramatic reductions in standards, the proposal was surprisingly brought forward by the CPBAO council with minimal external consultation or thorough public consideration of existing evidence addressing potential consequences. Moreover, psychologists themselves were underrepresented in the voting members (6 of 15, 40% representation) while non-psychologists played a predominant role in the decision-making process (9,10).

Given the sweeping scale of the proposal – if adopted, Ontario will become the largest jurisdiction with the lowest psychologist competency standards in North America – there has been a substantial public outcry in response to the proposed changes. More than 5000 individuals have signed a petition to halt these proposed changes (11). The Ontario Psychological Association has spoken out in opposition to the changes (12). Extensive grass-roots organization and advocacy have been undertaken, and the issue has been discussed extensively in the Canadian media (13), such as the *Globe and Mail* (14) and *Toronto Star* (15), and, internationally, in *The New York Times* (16). An advocacy group recently constructed a website to direct CPBAO members as well as members of the public for more information (17). Most recently, more than 1,300 individuals have signed an open letter to the CPBAO articulating concerns about these reduced competency standards (18).

In this contentious context, the goal of the current study was to characterize attitudes towards the proposed changes in a sample of general community adults in Ontario. Specifically, the study added questions to an ongoing longitudinal observational study, originally launched in 2018 to monitor the impacts of cannabis legalization. Over time, the cohort study has addressed not only impacts of cannabis legalization (19–22) but also expanded its scope to broader public health questions (23–27). Assessments are approximately every six months and items relating to the proposed changes were added to the wave running from October 17^th^ to November 17^th^. Participants were surveyed about the extent to which they approved or disapproved of the proposed changes with structured and open-ended follow-up queries about reasons for approval/disapproval. In addition to overall attitudes, a secondary goal was to characterize potential differences by population subgroups.

## METHODS

### Participants

The PATH CANN cohort has been described in detail in previous publications (21–29). Briefly, the original cohort comprised 1502 general community adults recruited from an existing research registry at St. Joseph’s Healthcare Hamilton the month before Canadian cannabis legalization in October 2018 and the current sample reflects *n*=1141 (76% retention since 2018). This includes the exclusion of one participant who did not meet the data quality control standards (i.e., 3+ correct answers on 5 embedded items with unambiguous responses, e.g., “*For this question, answer…*”). Demographic characteristics are in Table 1, and the participants can be generally characterized as middle-aged, employed adults; previous analyses have compared the cohort to provincial demographics and found it to be substantively similar (30), albeit with slightly higher education levels, somewhat lower racial diversity, and overrepresentation of females. All data were collected using REDCap electronic data capture (31). The protocol was approved by the Hamilton Integrated Research Ethics Board (#4699).

**Table 1.**
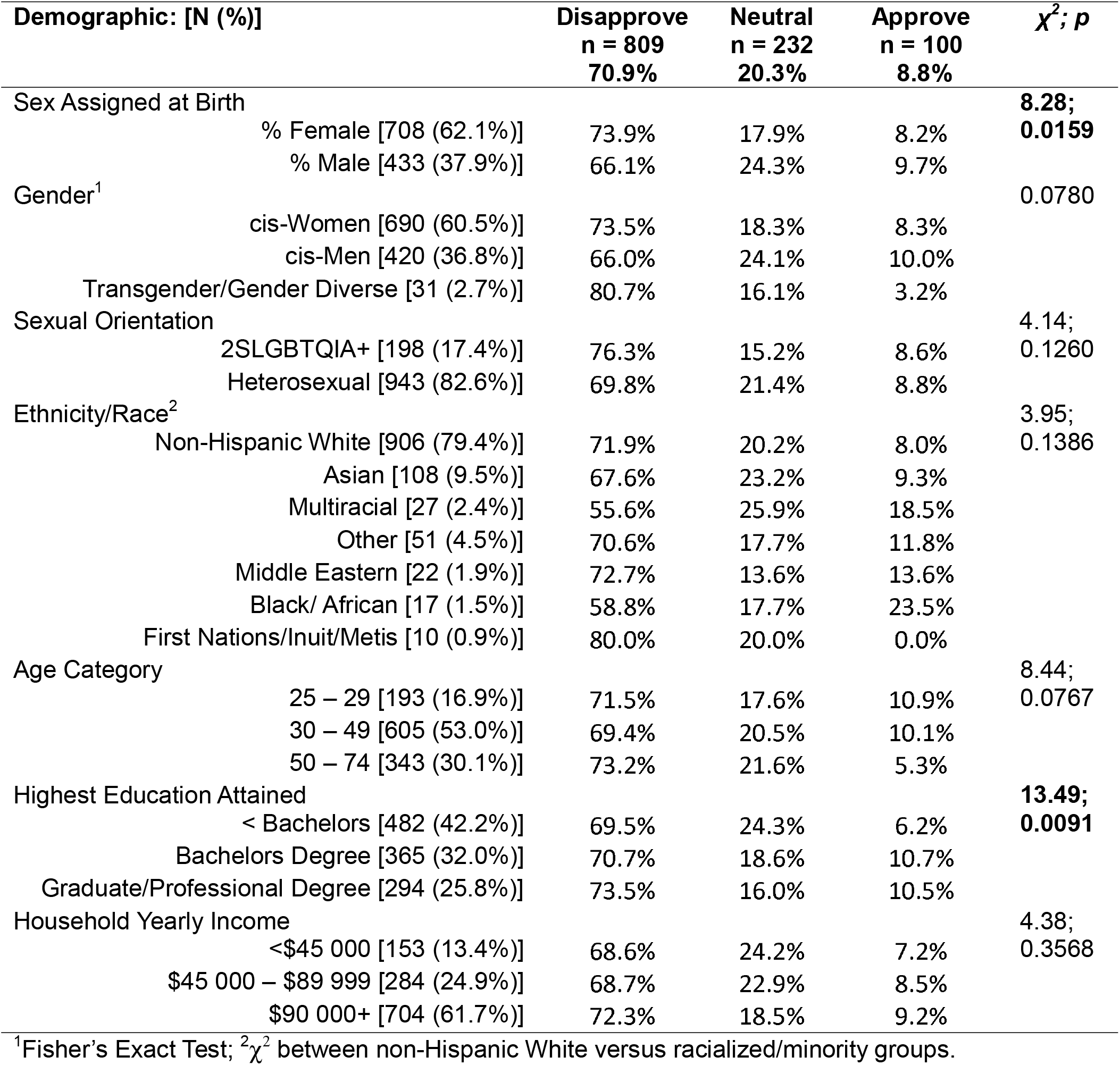
Demographics and demographic subgroup differences in disapproval/approval of the proposed changes to the psychologist competency standards.

### Assessment

The items assessing attitudes toward the proposed changes were purpose-built and based on the CPBAO description and the associated discussion. After a description of the proposed changes, participants rated their opinion with five response options (i.e., Strongly Disapprove, Somewhat Disapprove, Neither Approve Nor Disapprove, Somewhat Approve, Strongly Approve), with branching to reasons based on approval ratings (no branching for Neither Approve Nor Disapprove). Reasons for approving or disapproving were taken from the CPBAO material and a discussion of the potential changes in the Psychology Department at St Joseph’s Healthcare Hamilton. An open response “Other” was provided for additional reasons or comments. Reasons for approval/disapproval first permitted selecting all that applied followed by the same options but a forced choice for the most important reason for a participant’s perspective. The exact items are listed in the Supplementary Materials.

### Data Analysis

Descriptive statistics characterized the sample and overall opinion rates, followed by χ^2^ tests for differences by subgroups. Qualitative open response text was examined for alternative or diverging perspectives not captured by the response options.

## RESULTS

### Overall Preferences

As shown in Figure 1, the large majority disapproved of the proposed changes (71%), whereas 20% reported no opinion and 9% approved of the changes. Comparing gradations of disapproval and approval, there was asymmetric strong disapproval (44%; more common than somewhat disapproval) relative to strong approval (less common than approving somewhat, 2% overall). Of note, strong disapproval was the modal response overall.

**Figure 1.**
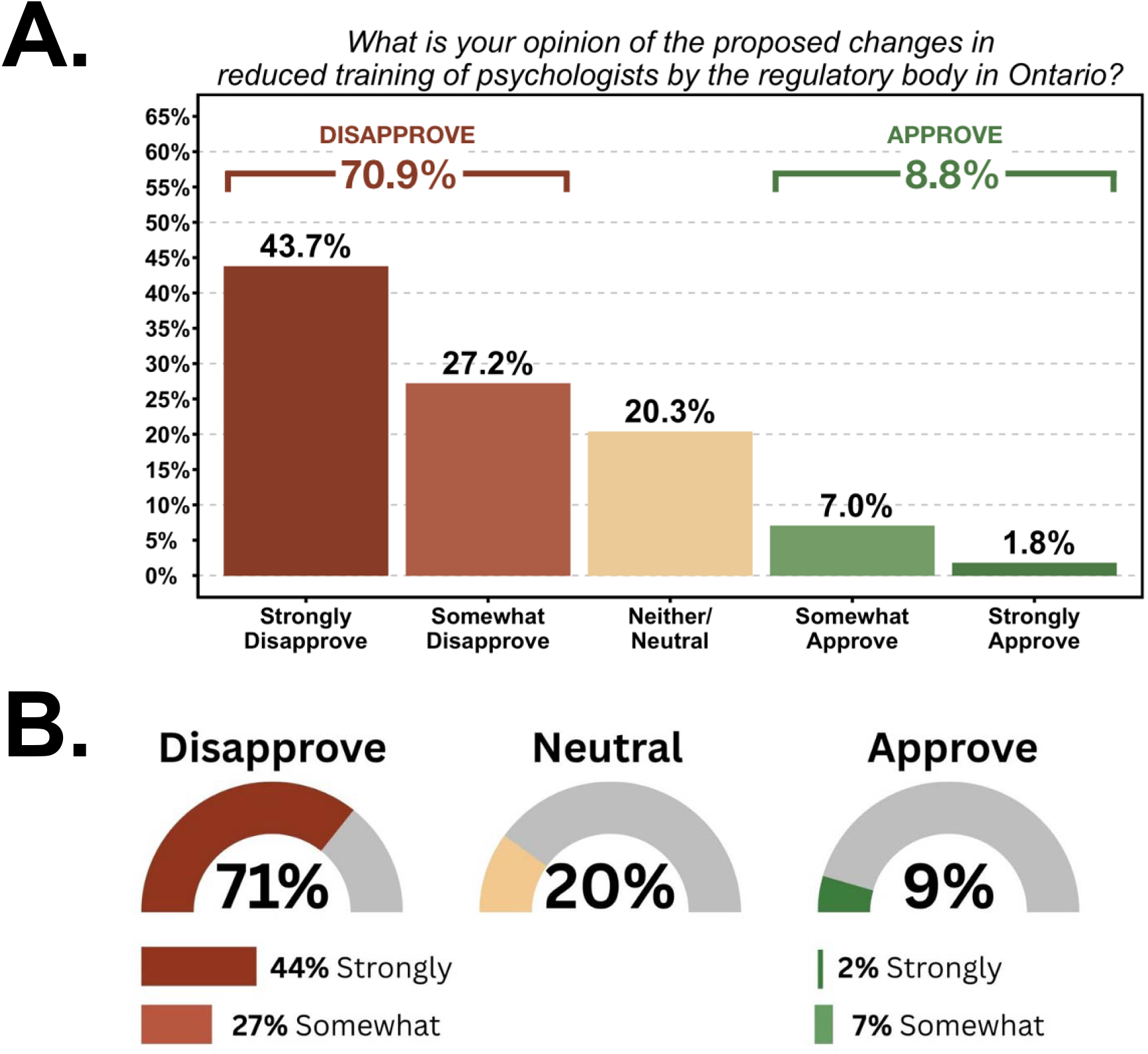
Overall perceptions of the proposed competency standards for psychologists in Ontario, Canada. Distinct visualizations are presented in Panels A and B for ease of interpretation.

### Reasons for Disapproval and Approval

Among non-mutually-exclusive options, the frequency of endorsement is in Figure 1. Nearly all participants were concerned that the changes would bring reduced quality of healthcare (94%) and large majorities were concerned there would be more healthcare providers, with a lower skillset and competencies (85%), a higher probability of misdiagnosis (78%), and increased risk in high-stakes contexts (69%). One third (33%) of participants endorsed all categories of adverse outcomes. Open field responses were given by 2.5% of participants and are excerpted in Table 2. Within the 9% who approved, the most common reason for approving was increased access to psychologists (86%), followed by increased numbers of psychologists (77%), followed by increased labour mobility (67%). Text responses are given in Table 2.

**Table 2.**
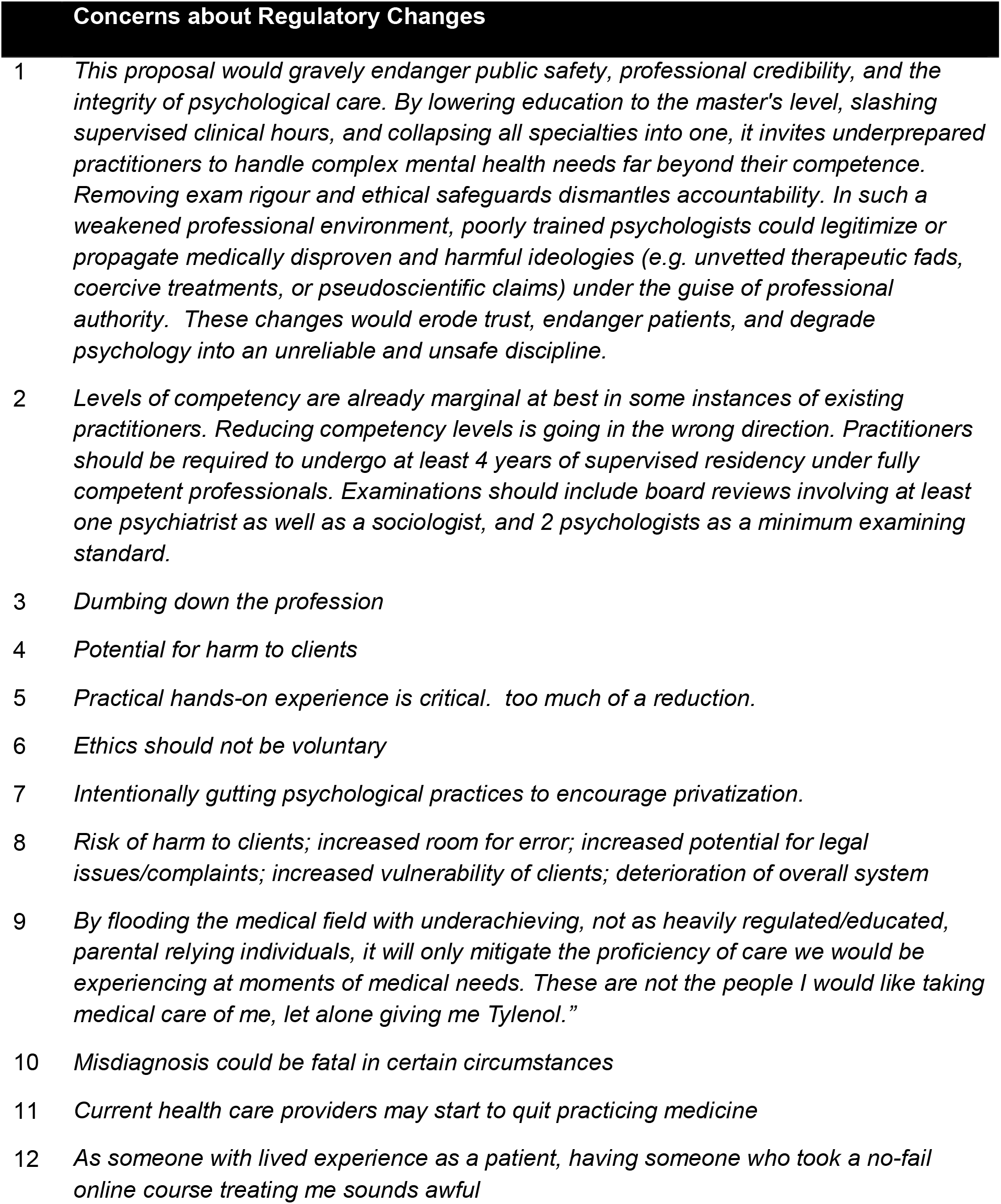
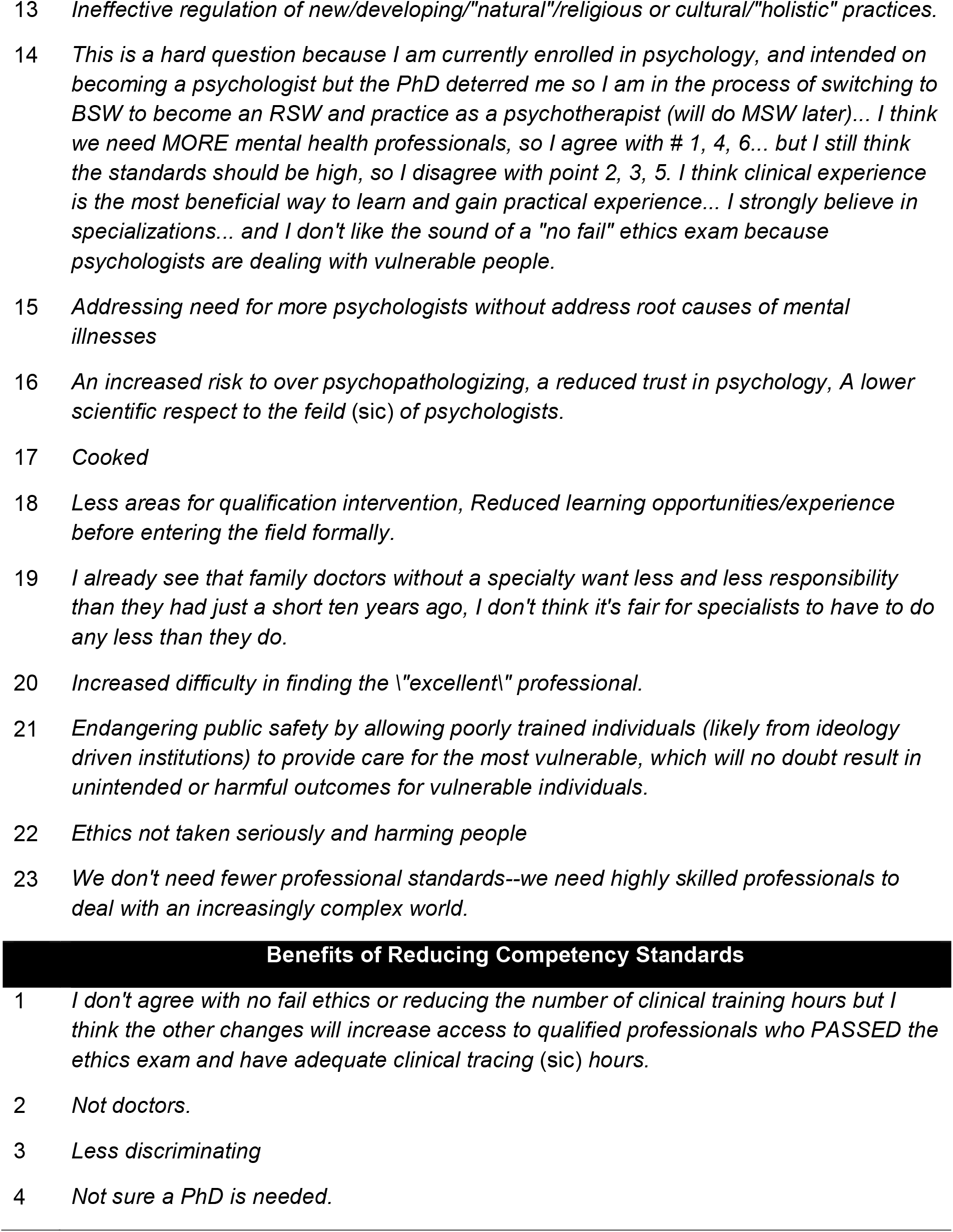
Open text responses for “Other” reasons for disapproving or approving for the changes.

In terms of the single most important concern, reduced quality of healthcare was chosen (62%). An increased number of healthcare providers with lower professional competency was chosen as the second biggest concern (22%). The third biggest concern chosen was an increased likelihood of misdiagnosis (9%) (Figure 2). Three participants responded “Other” as most important and their responses are excerpted in Table 2.

**Figure 2.**
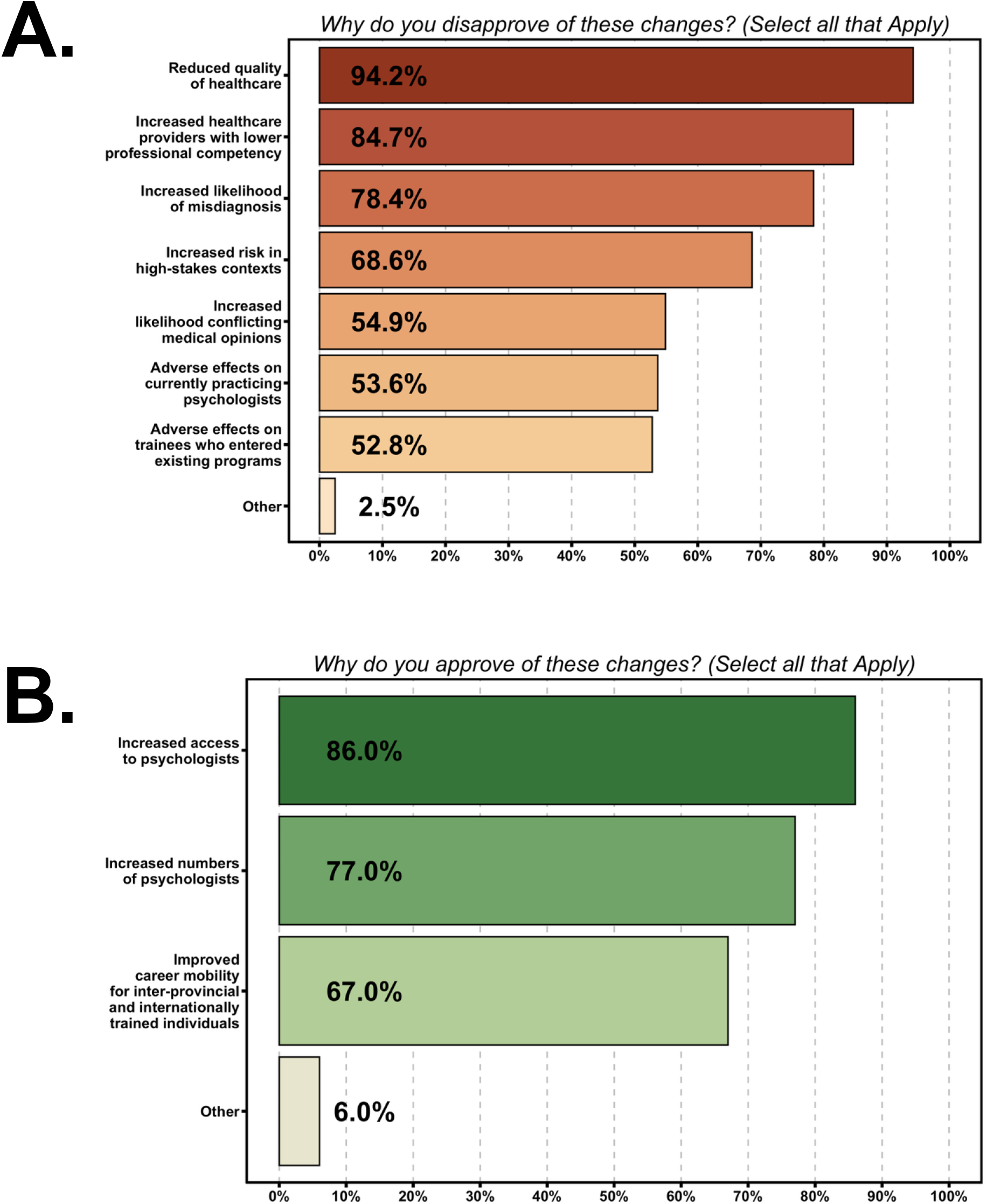
Reasons for disapproval and approval of the proposed changes. Response options were not mutually exclusive. Reasons for disapproval (n=809) are in Panel A and reasons for approving (n=100) are in Panel B.

In terms of the single most important reason for approving, increased access to psychologists was by far the most common (68%), followed by an increased number of psychologists (19%) and then labor mobility (11%) (Figure 2). Two participants responded “Other” as most important but did not provide text responses.

### Subgroup Differences

Subgroup differences are in Table 1. Sex differences were present such that female participants disapproved at a somewhat higher rate than male participants and educational differences were present such that lower education was associated with notably higher rates of Neither Approve Nor Dispprove. No significant differences were present for sexual orientation, race, education level, or income.

## DISCUSSION

The current study sought to characterize public opinion of the proposed reductions in competency standards for psychologists in Ontario, finding very strong evidence of high public disapproval. The overwhelming majority of the sample disapproved of the changes and the most common opinion was strong disapproval, approaching half of the total sample. In person-centred numbers, for every ten people, seven disapproved, two did not have an opinion, and one approved. Among participants who either disapproved or approved, there was an approximately eight times higher rate of disapproval relative to approval. Even more starkly, among participants with strong opinions (against or in favour), the rate of strong disapproval was 22 times higher than strong approval. Collectively, these data reveal very high public disapproval for the proposed changes in competency standards in the overall sample from any perspective on the data.

Among those who disapproved, more than 90% were concerned about reductions in competency standards degrading healthcare quality. This is consistent with Nova Scotia’s experience. After moving to a Master’s-level entry standard in 2011, the province observed that these providers did not meet the complex care needs of the population and in 2024 began seeking to reinstate the doctoral standard (32). According to the “*Assessment Submission for Entry-to-Practice Credential Change*” issued by the Nova Scotia Board of Examiners in Psychology (NSBEP), among other concerns they noted, “*Master’s graduates at the entry level have foundational skills in evidence-based assessment and treatment, but do not have sufficient knowledge and training to independently work with complex clients*.” (p. 21). The document provides perspective from a private practitioner on the NSBEP, “*We do not need more master’s-level psychologists. We need more doctoral-level psychologists ready to tackle complex cases from the outset. There is a reason accredited training programs are at a doctoral-level and require supervision by doctoral-level psychologists* (Appendix A; p. 46). Quebec similarly increased its requirements to the doctoral level in 2006 (33). Potentially related to this issue, the second most common public concern from the current sample was more providers with lower competency levels (∼85%). As an example of the importance of competent psychologists, a recent meta-analysis in forensic psychology reviewed 70 studies with over 55,000 offenders, finding that sexual, violent, and domestic recidivism was most reduced “*when programmes received consistent hands-on input from a qualified registered psychologist*” (p, 1) (34). It is also consistent with the substantially lower pass rates on the EPPP in jurisdictions that do not require doctoral level training (35).

At nearly 80%, the third most common reason for concern was misdiagnosis and the risks of misdiagnosis are indeed highly significant (36). This is especially true for complex cases such as individuals experiencing mental health crises, criminal justice-involved individuals, or those with neurodevelopmental or neurocognitive disorders. Lower training depth and less specialist training predicts higher diagnostic error and reduced treatment quality (34,37). Neuropsychologists are particularly needed in hospital settings, given the utility of their assessments and ability to diagnose complex psychiatric and neurocognitive disorders (38). In healthcare provider education, it is well documented that knowledge deficits are a major cause of diagnostic error, and individualized feedback on clinical diagnostic dispositions, as practiced in clinical supervision, has been shown to improve diagnostic abilities (39). Moreover, diagnostic errors in mental health care carry significant economic burden. For example, a US study found that patients with bipolar I who were misdiagnosed as having major depressive disorder incurred on average $6,500 per person in annual healthcare cost (40). A Canadian study estimated that psychological distress, absent an (appropriate) diagnosis, was associated with $3,364 per capita healthcare cost annually (41). These numbers do not even take into account the societal cost of absences, risks related to inappropriate medication regimens, decompensation leading to hospitalization, and greater utilization of psychiatric emergency services (42). More broadly, misdiagnosis poses a significant risk of stigmatization (43–45) and poses both risk to the patient but also fiduciary risk (e.g., legal liability, financial loss) when an individual or an organization fails to meet the standards of care owed to a patient.

The small proportion of the sample who approved of the changes predominantly reported doing so to increase access to psychological care (86%) and to increase the numbers of psychologists (77%). Improving inter-provincial mobility, one pretext for the proposed changes, was the least endorsed reason for approval.

In terms of subgroup differences, significant sex differences were present and so were differences in educational attainment. Although these were statistically significant, the absolute magnitudes of difference were relatively small and the more striking finding was that the pattern of preferences was, on the whole, highly consistent across population subgroups.

The current study had a number of strengths and limitations. Strengths include the large sample size and provision of empirical data to speak to a timely policy discussion, where no other direct data are available (to our knowledge). Limitations include that although this cohort is moderately representative of the population of Ontario (30), female and non-Hispanic White individuals were overrepresented, and it did not purposively ascertain a fully representative sample across the province. Similarly, the measure used was purpose-built and based on college documents and dialogue among professionals, not a psychometrically validated instrument. This limitation is mitigated by the fact that validated measures would not be expected to research questions like the one at hand. In this policy space, some empirical evidence is better than none, particularly when decisions to date have been guided largely by conjecture.

Collectively, the current study nonetheless addresses an important gap in knowledge, namely public sentiment and “voice” regarding the CPBAO’s proposed reductions in competency standards for psychologists in Ontario. The results unambiguously reveal strong disapproval, with the prepotent reasons being degraded healthcare quality, proliferation of lower competency professionals, increases in misdiagnosis, and greater risk in high-stakes contexts. The sentiment might best be captured in one participant’s qualitative comments: “*We don’t need fewer professional standards--we need highly skilled professionals to deal with an increasing complex world*.” Supporters of the changes were comparatively few and rarely expressed strong support. Although a critical exegesis of the pretexts for the proposed changes falls beyond the scope of this article, the findings suggest that, at least from the perspective of the general public, alternative approaches to increasing access to well-trained competent psychologists are warranted. Whether the CPBAO or healthcare policymakers will be responsive to these findings or the public cavalcade of negative reactions is an open question.

## Data Availability

All data produced in the present study are available upon reasonable request to the authors and approval of the research ethics board.

## Acknowledgements

The authors are grateful for the excellent research support of Ms. Jessica Gillard, Laura Lee, and Jane DeJesus. The authors are also grateful to Dr. Amanda Doggett for feedback on an earlier version of this manuscript. The PATH CANN longitudinal observational cohort study is funded by the Canadian Institutes of Health Research. JM is supported by the Peter Boris Chair in Translational Addiction Research and a Tier 1 Canada Research Chair in Translational Addiction Research (CRC-2020-00170).

## SUPPLEMENTARY MATERIALS

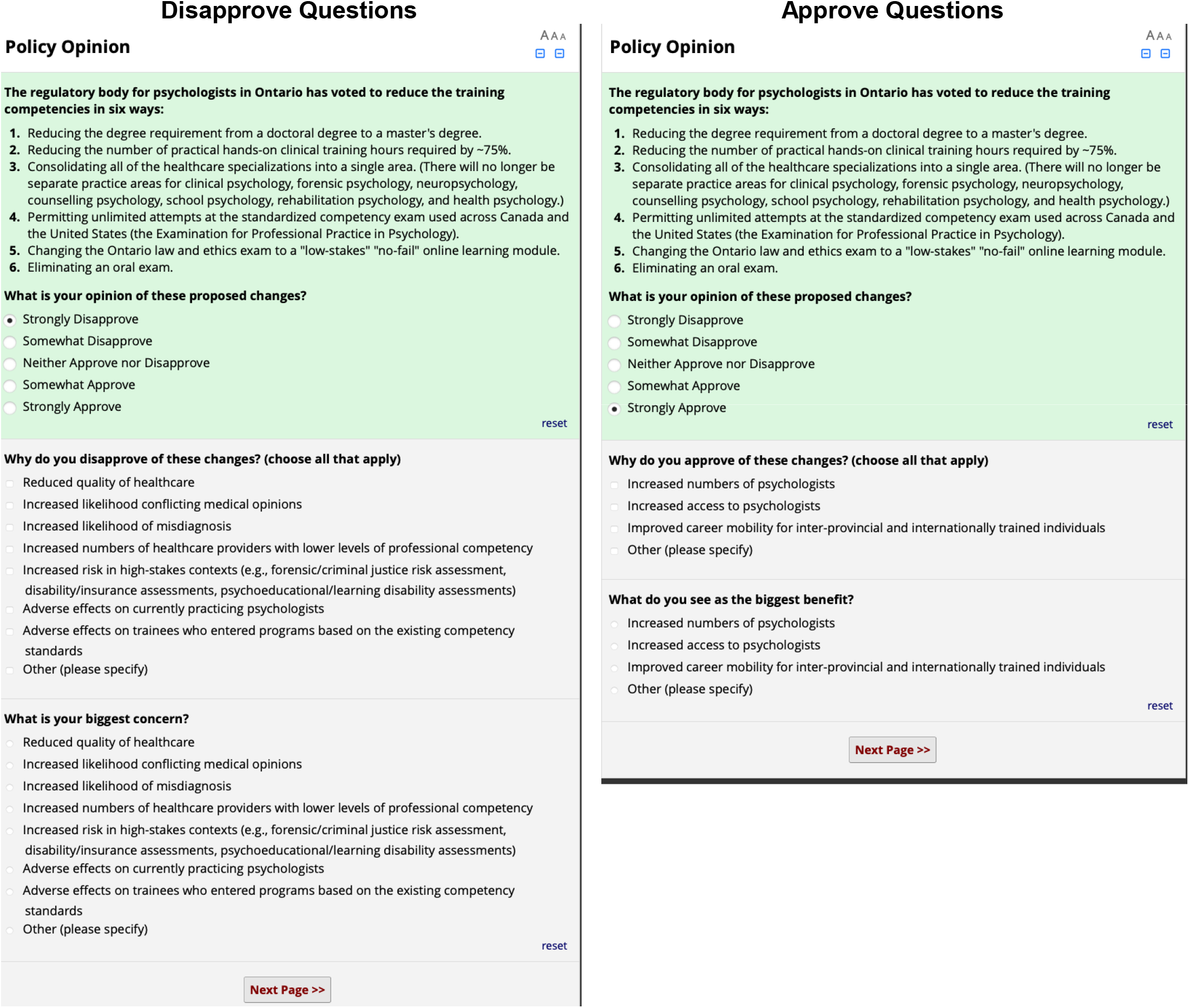

## Notes

### Competing Interest Statement

The authors have declared no competing interest.

### Author Declarations

This study was approved by the Hamilton Integrate Research Ethics Board.

